# A multi-omics study of circulating phospholipid markers of blood pressure

**DOI:** 10.1101/2020.12.08.20245050

**Authors:** Jun Liu, Paul S. de Vries, Fabiola Del Greco M., Åsa Johansson, Katharina E. Schraut, Caroline Hayward, Ko Willems van Dijk, Oscar. H. Franco, Andrew A. Hicks, Veronique Vitart, Igor Rudan, Harry Campbell, Ozren Polašek, Peter P. Pramstaller, James F. Wilson, Ulf Gyllensten, Cornelia M. van Duijn, Abbas Dehghan, Ayşe Demirkan

## Abstract

**Backgrounds:** High-throughput techniques allow us to measure a wide-range of phospholipids which can provide insight into the mechanisms of hypertension. We aimed to conduct an in-depth multi-omics study of various phospholipids with systolic blood pressure (SBP) and diastolic blood pressure (DBP).

**Methods:** The associations of blood pressure and 151 plasma phospholipids measured by electrospray ionization tandem mass spectrometry were performed by linear regression in five European cohorts (n = 2,786 in discovery and n = 1,185 in replication). We further explored the blood pressure-related phospholipids in Erasmus Rucphen Family (ERF) study by associating them with multiple cardiometabolic traits (linear regression) and predicting incident hypertension (Cox regression). Mendelian Randomization (MR) and phenome-wide association study (pheWAS) were also explored to further investigate these association results.

**Results:** We identified six phosphatidylethanolamines (PE 38:3, PE 38:4, PE 38:6, PE 40:4, PE 40:5 and PE 40:6) and two phosphatidylcholines (PC 32:1 and PC 40:5) which together predicted incident hypertension with an area under the curve (AUC) of 0.61. The identified eight phospholipids are strongly associated with triglycerides, obesity related traits (e.g. waist, waist-hip ratio, total fat percentage, BMI, lipid-lowering medication, and leptin), diabetes related traits (e.g. glucose, HOMA-IR and insulin) and prevalent type 2 diabetes. The genetic determinants of these phospholipids also associated with many lipoproteins, heart rate, pulse rate and blood cell counts. No significant association was identified by bi-directional MR approach.

**Conclusion:** We identified eight blood pressure-related circulating phospholipids that have a predictive value for incident hypertension. Our cross-omics analyses show that phospholipid metabolites in the circulation may yield insight into blood pressure regulation and raise a number of testable hypothesis for future research.

## Introduction

Long-term high blood pressure, of which 90-95% essential hypertension, is a major risk factor for cardiovascular diseases, e.g. coronary artery disease, stroke, heart failure, atrial fibrillation, etc^1^. Pervious study showed that the patients with essential hypertension have abnormal sodium-lithium counter transport across the red cell membrane, and that the level of transport is heritable^2^. Phosphatidylcholines (PC), phosphatidylethanolamines (PE), lysophosphatidylcholines (LPC), PE-based plasmalogens (PLPE), ceramides (CERs) and sphingomyelin (SPM) are groups of phospholipids that have a key function in the bilayer of (blood) cell membranes^3^. Although changes of membrane phospholipids in essential hypertension have been recognized and studied for a long time^4^, these previous studies either focused on animal models or overall phospholipid groups with limited resolution in the measurement. More detailed characterization of phospholipids in relation to hypertension at the population level is lacking.

In recent decades, high-throughput mass spectrometry (MS) has offered the opportunity to determine phospholipids on the chemical molecular level with high resolution at a low price. Thus, phospholipid panels with detailed characterisation are increasingly adopted in large epidemiological studies^5-11^. Despite these developments, the number of studies on the role of phospholipid profiles in hypertension and blood pressure is still limited. Very few studies of blood pressure and hypertension have investigated phospholipid profile in high resolution^12-15^. The study by Kulkarni et al. examined 319 (phospho)lipids in 1,192 Mexican-Americans^12^ and found that diacylglycerols (DG) in general and DG 16:0/22:5 and DG 16:0/22:6 in particular are significantly associated with systolic (SBP), diastolic (DBP) and mean arterial pressures as well as the risk of incident hypertension^12^. Stefan et al studied 135 cases and 981 non-cases of incident hypertension in a European study and identified four phospholipids and two amino acids which could improve the predictive performance of hypertension in addition to the known risk markers^15^. To our knowledge, no large-scale epidemiological study of blood pressure and/or hypertension with high-throughput measured phospholipids has been performed with replication in an independent study or has studied in detail the mechanism of the associations.

The aim of this study was to conduct an in-depth multi-omics study of the associations and causality of the associations of phospholipids with SBP and DBP, which are the diagnostic variables of hypertension, through metabolomics, genomics and phenomics. To this end, we investigated the association of blood pressure and 151 quantified phospholipids including 24 SPMs, 9 CERs, 57 PCs, 15 LPCs, 27 PEs, and 19 PLPEs, in 3,971 individuals from five European populations. Using Mendelian Randomization (MR), we further investigated the causality in these relationships. The potential genetic pleiotropy between phospholipids and blood pressure was also explored.

## Methods

### Population description

This study was conducted using five populations throughout Europe. The individuals with both blood pressure and phospholipid measure available were included: (1) the CROATIA-Vis study conducted on the island of Vis, Croatia (n = 710)^16^, (2) the Erasmus Rucphen Family (ERF) study, conducted in the Netherlands (n = 717)^17^, (3) the Northern Swedish Population Health Survey (NSPHS) in Norrbotten, Sweden (n = 678)^18^, (4) the Orkney Complex Disease Study (ORCADES) in Scotland (n = 681)^19^, and finally (5) the MICROS study from the South Tyrol region in Italy (n = 1,185)^20^ which was included for replication. Fasting blood samples were collected for the biochemical measurements. All studies were approved by the local ethics committees and all participants gave their informed consent in writing.

The association tests of phospholipids and blood pressure were performed on the same baseline data, for each of the five studies. The predictive analysis was performed in ERF study in which we collected follow-up data from March 2015 to May 2016 (9-14 years after baseline visit). During the follow-up, a total of 572 participants’ records from the 717 individuals included in the baseline analysis were scanned for common diseases in general practitioner’s databases.

### Phospholipids measurements

As part of the European Special Populations Research Network (EUROSPAN) project, the absolute concentrations (µM) of 151 lipid traits in plasma were centrally measured by electrospray ionization tandem mass spectrometry (ESIMS/MS), including 24 SPMs, 9 CERs, 57 PCs, 15 LPCs, 27 PEs and 19 PLPEs. The methods used have been validated and described previously^21,22^. For each lipid molecule, we adopted the naming system where lipid side chain composition is abbreviated as x:y, where x denotes the number of carbons in the side chain and y the number of double bonds. For example, PC 34:4 denotes an acyl-acyl PC with 34 carbons in the two fatty acid side chains containing four double bonds.

### Covariates

Supplementary Table 1 describes how SBP and DBP, T2D status, total cholesterol (TC), HDL-C, lipid-lowering medication usage, body mass index (BMI) and antihypertensive medication are measured or defined in the cohorts. In all cohorts, blood pressure was measured by automated reading in the sitting position after a rest. The medication information was collected during the personal interview. For the additional analysis in ERF only (described below), we imputed the missing values by multiple imputation in R package ‘mice’ and followed the Rubin’s rules^23^.

### Discovery and replication analysis

Lipids were natural log-transformed and standardized (mean-centered and divided by their standard deviation). We corrected blood pressure levels for antihypertensive medication use by adding 15 mmHg to the SBP and 10 mmHg to the DBP of users of antihypertensive medication^24-28^. As all of the five cohorts included closely related individuals, family relationship based on the genotype was adjusted for in the analysis by extracting polygenic residuals for the phenotypic traits, by using the polygenic option in GenABEL package in R^29^.

In each study, we used linear regression to examine the association between each of the phospholipids and blood pressure individually. Blood pressure variables were used as dependent variables and phospholipids were used as independent variables. We performed a discovery analysis in CROATIA-Vis, ERF, NSPHS, and ORCADES, adjusting for age and sex (model 1). Results from the four discovery populations were meta-analyzed with inverse-variance weighted fixed-effects model using the METAL software^30^. To correct for multiple testing, we used Bonferroni correction using the number of 70 independent components extracted from the 151 directly measured phospholipids (P-value < 7.1 × 10^−4^, 0.05/70). Matrix Spectral Decomposition was separately used to calculate the number of independent equivalents^31^ in each of the four discovery studies. Bonferroni correction was done for 70 tests which was the highest number obtained in CROATIA-Vis. We did not correct for the number of blood pressure variables as SBP and DBP are highly correlated (R = 0.65, P-value < 2.2 × 10^−16^ in ERF, n = 2,802).

We replicated our findings in MICROS (n = 1,185) using the same statistical framework as in the discovery analysis and using a Bonferroni correction for the independent number of tested associations, i.e., equivalents of the significant phospholipids.

In a combination of all five cohorts, we examined a further model (model 2) to assess the impact of potential confounders and mediators by additionally adjusting for BMI, HDL-C, TC, lipid-lowering medication and type 2 diabetes (T2D) status. We checked the pairwise Pearson’s correlation matrices of the blood pressure related phospholipids in ERF adjusting for age, sex and family relationships.

### Association with hypertension and cardiometabolic traits

The phospholipids significantly associated with SBP or DBP were tested for the association with the occurrence of hypertension during the follow-up in ERF. The incident cases were defined as the participants free of hypertension at baseline who were diagnosed with hypertension at follow-up by general practitioners. Time-to-event was defined as the time from the enrollment date at baseline to either the onset date of disease, date of death, date of censoring (moving away) or date of follow-up collection. Cox proportional regression analysis was used to evaluate the individual effect of phospholipids considering of the follow-up time (time-to-event). To determine the joint effect of the phospholipids on the discrimination of future hypertension patients, we calculated the area under the receiver operator characteristics (ROC) curve (AUC). We further determined whether the addition of the listed phospholipids increase the AUC value of the factors in the Framingham risk score for hypertension which includes age, sex, SBP, DBP, BMI, cigarette smoking and parental hypertension (Framingham model)^32^. Integrated Discrimination Improvement test (IDI) and continuous Net Reclassification Improvement test (NRI) were performed to compare different joint models.

In ERF, we further examined the association of these identified phospholipids with known cardiometabolic traits, including adiponectin, albumin, alcohol consumption, anti-diabetic and anti-hypertensive medications, BMI, creatinine, C-reactive protein (CRP), glucose, HDL-C, insulin, intima-media thickness, LDL-cholesterol, heart rate, homeostatic model assessment-insulin resistance (HOMA-IR), leptin, lipid-lowering medication, metabolic syndrome, plaque score, pulse wave velocity (PWV), resistin, smoking status, TC, total fat percentage, triglycerides, T2D, waist circumference and waist-to-hip ratio. The description and measurement methods of the above mentioned cardiometabolic traits can be found in our previous reports^33-38^. The distributions of adiponectin, insulin, leptin, triglycerides, CRP, HOMA-IR and resistin are skewed and therefore were log-transformed before performing the analysis. We used the standardized residuals of natural-log-transformed phospholipid levels as the dependent variable, adjusted for age, sex and family relationship. A hierarchical clustering approach was used to cluster the cardiometabolic traits^39^. We estimated the false discovery rate (FDR < 0.05) by Benjamini & Hochberg method considering of the gathering of categorical and continuous variables.

### Mendelian Randomization and phenome-wide association study

MR is a statistical method which uses the effect of genetic variants determining an exposure and test its association with the outcome under study, based on the assumption that the genetic variant is inherited independent of the confounding variables^40^. We performed a two-sample bi-directional MR of the 11 significant associations of phospholipids and SBP or DBP. We used summary statistics level data of blood pressure and phospholipids^7,28^ utilizing the pipeline in the R-package *TwoSampleMR*^41^. In brief, the genetic instrument was based on the top genetic determinant SNPs with linkage disequilibrium (LD) R^2^ < 0.05 within 500kbps clumping distance. The proportion of variance in the exposure explained by the genetic variance (R^2^) and F statistics were calculated to estimate the statistic power of MR. As the sample size in the phospholipid GWAS in the ESIMS/MS platform is small (n = 4,034), to increase the explained variance of the instrumental variable, P-value < 1.0 × 10^−7^ was used to define the genetic determinants of phospholipids. The GWAS summary statistics of the same phospholipids available in Biocrates metabolomics platform were also used additionally to increase the statistical power (n = 7,478)^42^. For genetic determinants of blood pressure, the genome-wide significance level (P-value < 5 ×10^−8^) was used. Inverse-variance weighted MR was used with weighted median, sample mode and weighted mode methods as sensitivity to investigate pleiotropy. MR-Egger regression was used to control the directional horizontal pleiotropy, and the Egger estimates on the intercept was used for the heterogeneity tests^43,44^. The Bonferroni corrected P-value with independent equivalents of phospholipids was used as the significance level.

### Phenome-wide association study (pheWAS)

We further studied the pleiotropic effect of the genetic determinants of the identified phospholipids using phenome-wide association study (pheWAS) by data-mining from previous publications^45^. For the top SNPs of either phospholipids used in MR, we looked up their pheno-wide associations in GWAS ATLAS^45^. We estimated the false discovery rate (FDR < 0.05) by Benjamini & Hochberg method.

## Results

### Association analysis

Baseline characteristics of the five participating cohorts are shown in Table 1. The mean age ranged from 47.1 (with standard deviation 20.8) years old in NSPHS to 56.6 (with standard deviation 15.6) years old in CROATIA-Vis. and the proportion of females ranged from 53.2 % in NSPHS to 57.0% in ORCADES. The means and standard deviations of the concentration of the 151 phospholipids across the five cohorts are shown in Supplementary Table 2 and Supplementary Figure 1. Most of the phospholipids have similar concentrations across cohorts, except for PLPE 18:1, PLPE 18:0 and PLPE 16:0. The associations of all 151 phospholipids with blood pressure in the discovery panel, replication panel and combination are shown in in Supplementary Table 3. Volcano plots in Figures 1A and 1B show the meta-analysis results of the discovery panel in a J shape. Five phospholipids (PC 32:1, PC 40:5, PE 38:4, PE 40:5 and PE 40:6) were significantly associated with SBP, and seven phospholipids (PE 38:3, PE 38:4, PE 38:6, PE 40:4, PE 40:5, PE 40:6 and LPC 22:4) were associated with DBP using Bonferroni corrected significance threshold. For the significant phospholipids found in the discovery analysis, only LPC 22:4 was associated inversely to DBP. Eleven of the 12 significant associations from the discovery were replicated in MICROS based on the following adjusted P-value thresholds: 0.017 for SBP and 0.013 for DBP (Figure 1). Only LPC 22:4 did not replicate in MICROS. Further, we focused on the 11 significant associations with eight unique phospholipids which were replicated.

**Table 1.** Baseline characteristics of the study population in the association analysis.

|  | Discovery |  |  |  | Replication |
| --- | --- | --- | --- | --- | --- |
|  | CROATIA-Vis | ERF | NSPHS | ORCADES | MICROS |
| N | 710 | 717 | 678 | 681 | 1,185 |
| Age (y) | 56.6 (15.6) | 51.9 (14.2) | 47.1 (20.8) | 57.0 (13.9) | 45.7 (16.4) |
| Sex (% women) | 57.40 | 59.00 | 53.2 | 57.9 | 56.0 |
| Body mass index (kg/m<sup>2</sup>) | 27.4(4.3) | 27.0 (4.4) | 26.4 (4.8) | 27.7 (4.8) | 25.6 (4.8) |
| HDL-C (mmol/L) | 1.10(0.16) | 1.29 (0.36) | 1.60 (0.41) | 1.57 (0.42) | 1.68 (0.38) |
| TC (mmol/L) | 5.12(0.99) | 5.67 (1.08) | 5.86 (1.33) | 5.56 (1.14) | 5.87 (1.19) |
| Lipid-lowering medication use (%) | 3.00 | 16.60 | 10.47 | 4.10 | 5.47 |
| Systolic blood pressure (mmHg) | 137.8 (24.45) | 141.7 (22.0) | 122.8 (18.6) | 130.1 (18.46) | 132.6 (20.5) |
| Diastolic blood pressure (mmHg) | 80.5 (11.47) | 80.6 (10.24) | 74.1 (7.8) | 76.0 (9.6) | 79.6 (11.3) |
| Antihypertensive medication use (%) | 24.1 | 25.2 | 20.5 | 39.1 | 14.6 |
| Type 2 diabetes (%) | 4.2 | 6.0 | 4.1 | 2.8 | 3.1 |

Values are mean (SD) or percentages. N refers to the largest sample size used in this study.

**Figure 1.**
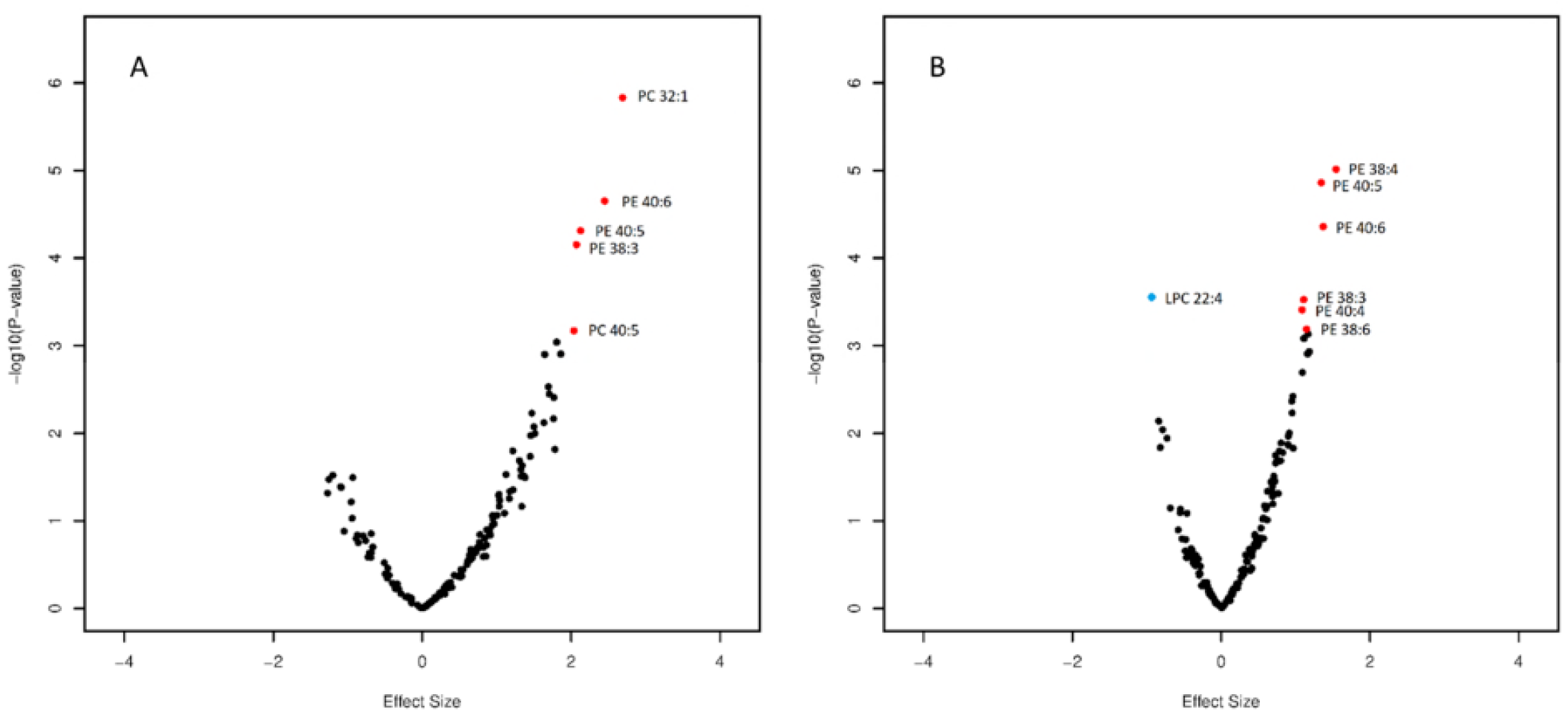
Association of phospholipids and blood pressure in model 1. Figure 1A: phospholipids associated with SBP. Figure 1B: phospholipids associated with DBP. Age, sex and family relationship were adjusted for in the regression analysis. Red: Lipids are significantly associated with blood pressure and replicated. Blue: lipid LPC 22:4 significantly associated with blood pressure but failed in the replication.

The significant associations were generally attenuated upon adjustment for BMI, HDL-C, TC, lipid-lowering medication and T2D status in model 2, with the proportion of the effect estimate decreased ranged from 4.5% for the association between PC 32:1 and SBP to 46.2% for the association between PE 40:6 and DBP, but all of the associations remained significant (Table 2). All the identified phospholipids were highly correlated with each other, while the correlation among the PEs was obviously higher than with the PCs or between the PCs (Supplementary Figure 2).

**Table 2.** Effect of adjustments on the association between selected lipids and blood pressure.

| Name | Model 1 |  |  |  |  |  | Model 2 |  |
| --- | --- | --- | --- | --- | --- | --- | --- | --- |
|  | Discovery<br>(N = 2,786) |  | Replication<br>(N = 1,185) |  | Combined<br>(N = 3,971) |  | Combined<br>(N = 3,937) |  |
|  | Effect | P-value | Effect | P-value | Effect | P-value | Effect | P-value |
| *Systolic blood pressure* |  |  |  |  |  |  |  |  |
| PC 32:1 | 2.7 | $1.5 \times 10^{-06}$ | 1.8 | $2.4 \times 10^{-04}$ | 2.2 | $2.6 \times 10^{-09}$ | 2.1 | $9.6 \times 10^{-08}$ |
| PC 40:5 | 2.0 | $6.7 \times 10^{-04}$ | 2.0 | $1.0 \times 10^{-04}$ | 2.2 | $3.2 \times 10^{-09}$ | 1.6 | $2.7 \times 10^{-05}$ |
| PE 38:3 | 2.1 | $7.0 \times 10^{-05}$ | 2.2 | $2.6 \times 10^{-05}$ | 2.1 | $6.8 \times 10^{-09}$ | 1.5 | $4.0 \times 10^{-05}$ |
| PE 40:5 | 2.1 | $4.9 \times 10^{-05}$ | 2.3 | $1.7 \times 10^{-05}$ | 2.2 | $3.2 \times 10^{-09}$ | 1.6 | $2.7 \times 10^{-05}$ |
| PE 40:6 | 2.4 | $2.2 \times 10^{-05}$ | 1.7 | $6.7 \times 10^{-04}$ | 2.0 | $8.6 \times 10^{-08}$ | 1.2 | $1.9 \times 10^{-03}$ |
| *Diastolic blood pressure* |  |  |  |  |  |  |  |  |
| PE 38:4 | 1.5 | $9.7 \times 10^{-06}$ | 1.2 | $7.6 \times 10^{-07}$ | 1.3 | $3.7 \times 10^{-11}$ | 0.9 | $1.3 \times 10^{-05}$ |
| PE 40:5 | 1.3 | $1.4 \times 10^{-05}$ | 1.4 | $6.7 \times 10^{-08}$ | 1.4 | $3.5 \times 10^{-12}$ | 1.0 | $3.4 \times 10^{-06}$ |
| PE 40:6 | 1.4 | $4.4 \times 10^{-05}$ | 1.3 | $3.5 \times 10^{-07}$ | 1.3 | $5.7 \times 10^{-11}$ | 0.7 | $3.4 \times 10^{-04}$ |
| PE 38:3 | 1.1 | $3.0 \times 10^{-04}$ | 1.3 | $1.8 \times 10^{-07}$ | 1.2 | $7.0 \times 10^{-10}$ | 0.8 | $4.0 \times 10^{-05}$ |
| PE 38:6 | 1.1 | $6.5 \times 10^{-04}$ | 0.8 | $2.5 \times 10^{-03}$ | 0.9 | $1.2 \times 10^{-05}$ | 0.6 | $2.7 \times 10^{-03}$ |
| PE 40:4 | 1.1 | $3.9 \times 10^{-04}$ | 1.9 | $3.1 \times 10^{-11}$ | 1.6 | $3.8 \times 10^{-13}$ | 1.2 | $7.2 \times 10^{-08}$ |

### Association with future hypertension and cardiometabolic traits

We studied the relationship between the identified phospholipids and incident hypertension in 447 available participants from the ERF study, including 92 patients with incident hypertension. None of the eight identified phospholipids (six PEs and two PCs) were individually significantly associated with incident hypertension in our study (Supplementary Table 4). But the joint effect of the eight phospholipids was significantly associated with incident hypertension (P-value = 5.0 × 10^−8^, Figure 2). Although in the phospholipids-only model, the discrimination between those with and without future hypertension is limited (AUC=0.61) and significantly lower than that of the Framingham model, adding the eight phospholipids significantly improved the AUC on top of the Framingham model from 0.75 to 0.76 (P_IDI_ = 0.02, P_NRI_ = 0.06, Figure 2).

**Figure 2.**
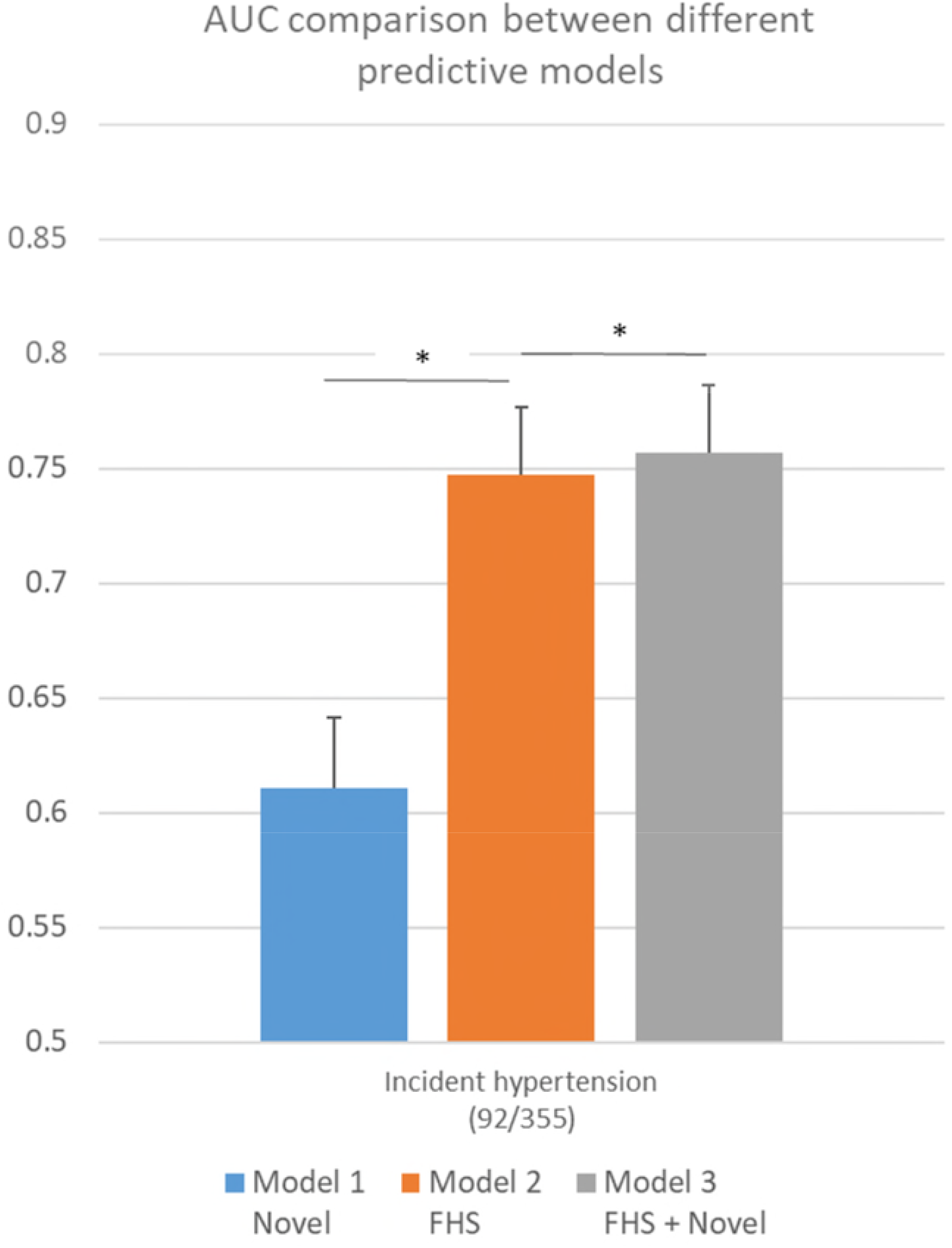
AUC comparison between eight phospholipids associated with either SBP or DBP, the factors included in the Framingham risk score and their combination. Model 1 *Novel*: The model includes phospholipids associated with either SBP or DBP only: PC 32:1, PC 40:5, PE 38:3, PE 40:6, PE 40:5, PE 38:4, PE 38:6, PE 40:4. Model 2 *FHS*: The model includes the factors from the Framingham risk scores of incident hypertension: age, sex, SBP, DBP, BMI, cigarette smoking and parental hypertension. Model 3 *FHS + Novel*: the advanced model adding factors in model 1 and Model 2. * P_IDI_ < 0.05. IDI: Integrated Discrimination Improvement test.

Figure 3 shows the association of the blood pressure-related phospholipids with the classical/clinical cardiometabolic traits measured in ERF (N = 818 analytical sample size). Triglycerides were strongly associated with all of the eight blood pressure-related phospholipids and form the first cluster themselves. Although the direction and strength of association are very similar to triglycerides, the association of triglycerides appears to be independent of the second cluster. The second cluster involved waist, waist-hip ratio, glucose, total fat percentage, BMI, leptin, use of anti-hypertensives, T2D, lipid-lowering medication, CRP, HOMA-IR, insulin and TC. Most of the significant associations were in the same (positive) direction of the association between blood pressure and related phospholipids. The third cluster had much fewer significant associations. We found associations between PCs and environmental exposures such as smoking and alcohol intake, but also heart rate, albumin, HDL and LDL-C and adiponectin. No significant association was found between the phospholipids and vascular-related variables, including PWV, intima-media thickness and plaque score.

**Figure 3.**
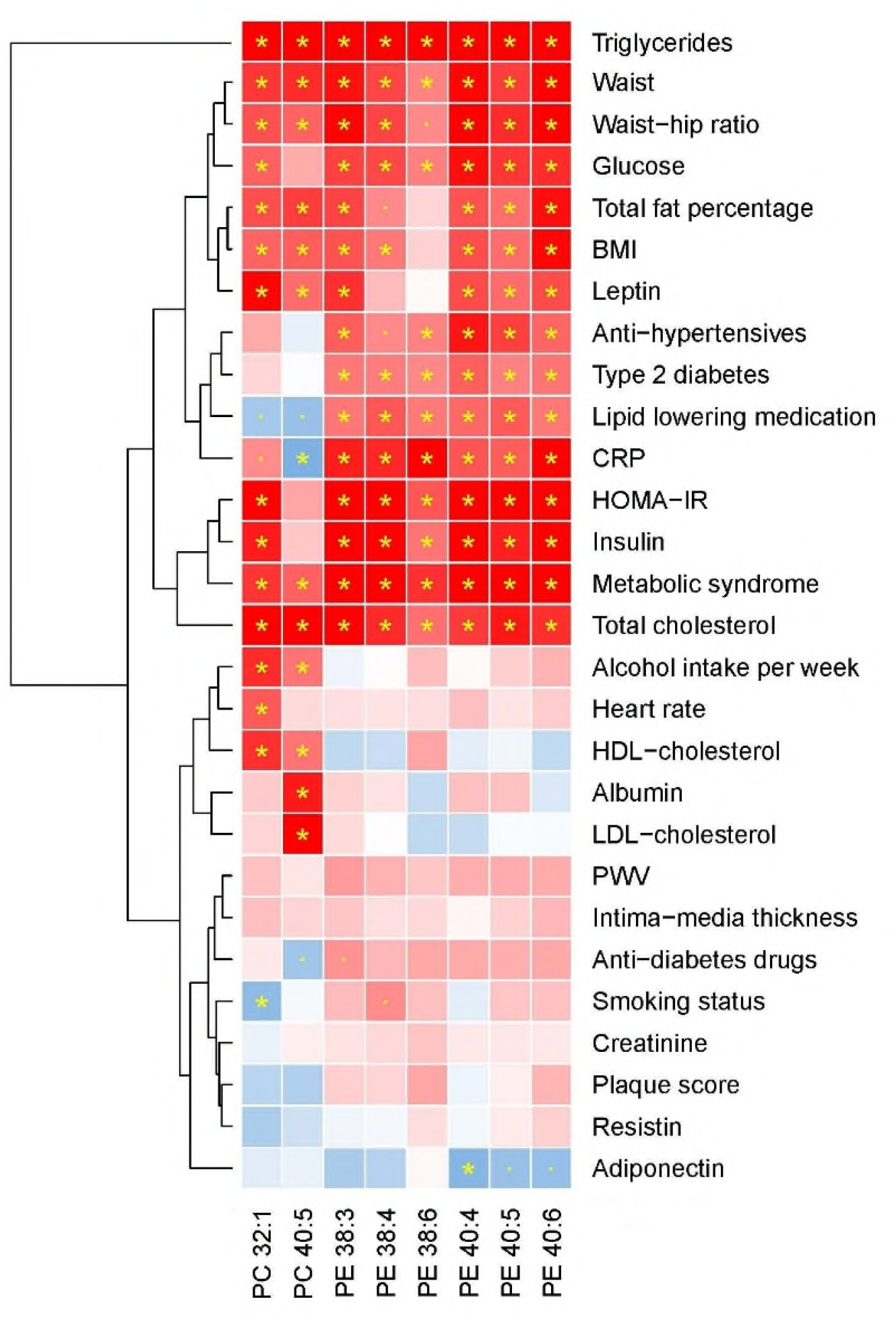
Association of blood pressure related phospholipids and cardiometabolic traits in ERF. Hierarchical clustering approach was used for the clustering. Red: positive association; blue: negative association. The depth of the color displays the strength of z score in the regression. * FDR < 0.05. . P-value < 0.05.

### Mendelian randomization and phenome-wide association study

The MR pipeline resulted in two to six independent SNPs included in the genetic risk score as instrumental variables for each phospholipid (R^2^ range from 2.7% to 5.2%), 471 SNPs for SBP (R^2^ = 4.0%) and 506 SNPs for DBP (4.3%). The F-statistics ranged from 55.1 for the MR in PE 40:4 to DBP to 105.7 in PE 40:5 to SBP and DBP. The two PCs, i.e. PC 32:1 and PC 40:5 were also performed using the summary statistics of Biocrates platform. However, no significant results were found in either MR test (Supplementary Table 5).

The top SNPs which were associated at genome-wide significance with blood pressure related phospholipids are rs174576, rs10468017, rs261338, rs12439649, rs740006 and rs7337573 and located in the protein-coding genes *TMEM258, FADS2, ALDH1A2, LIPC*, and antisense gene *RP11-355N15*.*1*, after considering for the linkage disequilibrium. In total, 1513 SNP-trait associations were identified from the pheWAS database^45^ after controlling for false discovery rate (Supplementary Table 6). The most highly significant related traits were in metabolic domain which are mainly lipoproteins, and blood cell counts. The next highly related traits are heart rate and pulse rate in the cardiovascular domain. Other highly significant related traits including male pattern baldness, height, glucose, etc. (Figure 4).

**Figure 4.**
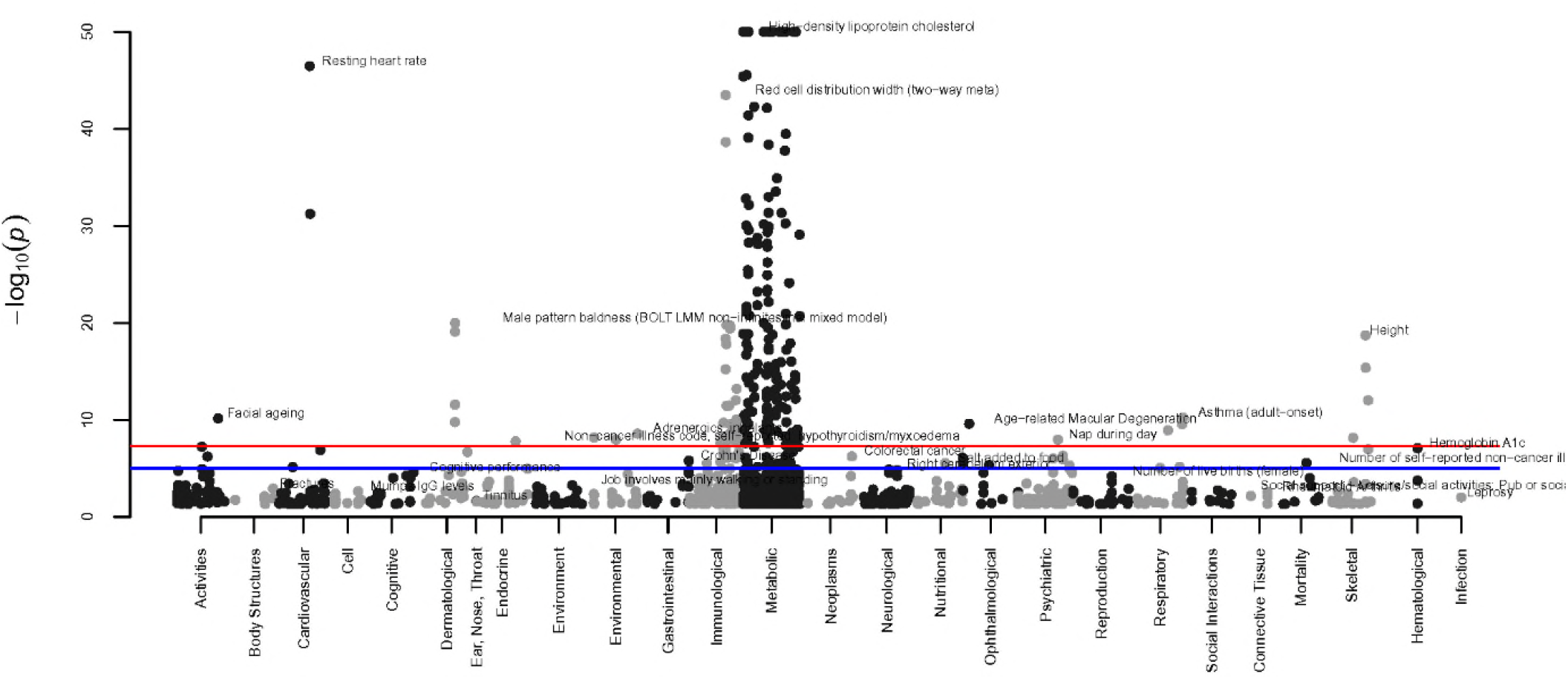
Results of the phenome-wide association study of the genetic determinants of the blood pressure related phospholipids. In each domain (x-axis), the top trait was annotated in the figure. The traits with P-value level less than 1.0 × 10^−50^ were annotated as 1.0 × 10^−50^ in the figure.

## Discussion

The current study identified and replicated the association of eight phospholipids with either SBP or DBP. These phospholipids jointly associated with incident hypertension and improved the discrimination model of incident hypertension. Strong associations were identified of these phospholipids with triglycerides, but also with obesity related traits (e.g., waist, waist-hip ratio, total fat percentage, BMI and leptin), T2D and related traits (e.g. glucose, HOMA-IR and insulin). Meanwhile, the genetic determinants of these phospholipids also highly and genome-widely associated with lipoproteins, blood cell counts, heart rate, pulse rate, glucose and many potential pleiotropic traits, e.g. male pattern baldness, height, etc. No significant association was identified between the genetic susceptibility of blood pressure and phospholipids by MR approach, in either direction.

We found a predictive effect of the joint phospholipids on future hypertension, which was consistent with the associations of these phospholipids and incident hypertension in the Mexican-American population^12^; among the 11 replicated associations in the current study, ten associations were replicated in the Mexican-American population using the current Bonferroni P-value adjustment (0.017 for SBP and 0.013 for DBP, Supplementary Table 7). Moreover, PC 40:5, PE 38:3, PE 38:4, PE 38:6, PE 40:5 and PE 40:6 were also associated with incident hypertension in their study. The similarity of the findings in populations of different ancestries suggests a probability of a generalizable biological process. This is in line with our finding that the significant associations of the identified phospholipids and blood pressure remain after adjustment for various potential confounders or mediators, suggesting that the associations are independent of these covariates. The association of the phospholipids with incident hypertension and the improved predictive performance with adding these phospholipids onto the Framingham model also implies that the phospholipid level may be a predictor of the occurrence of diagnosed hypertension. However, we could not confirm any causation by current MR approach. Further research is required to investigate this hypothesis.

The identified phospholipids are strongly associated with triglycerides, which share 1,2-diglyceride with PCs and PEs as a substrate in their biosynthesis^46^. This is in line with the previous finding that blood pressure is related to DG in general and DG 16:0/22:5 and DG 16:0/22:6^12^. We also found that DBP is related to PE 38:6 which also includes a fraction of PE 16:0/22:6. These blood pressure related phospholipids are also associated with obesity and diabetes related traits (Figure 3). This is consistent with our results that after adjustment for BMI, HDL-C, TC, lipid-lowering medication and T2D status in model 2, the effect estimate generally attenuated. It implies that these factors may act as mediators in the associations. However, as the associations were still statistically significant, we could not completely exclude the direct association of the abnormal phospholipid levels and blood pressure. A one sample based formal causal mediation analysis in a large-scale cohort is suggested to confirm their mediating effect on the association of these phospholipids and blood pressure.

All the blood pressure related phospholipids identified in the current study have side chains that are unsaturated fatty acids. The highly unsaturated fatty acids in the circulation are from dietary intake as humans do not have the enzymes to synthesize them. The circulating fatty acid levels are thus determined by both dietary intake and degradation, while the individual capability of degradation is partially determined by heritability^47^. If the degradation is abnormal, this will cause high level of exogenous unsaturated fatty acids in circulation, which subsequently leads to high level of phospholipids which contain these unsaturated fatty acid chains. This is consistent with the J shape (Figure 1) of the associations between blood pressure and phospholipids in fasting blood samples, most of which have a positive direction. A recent study reported a higher heritability in the phosphatidylcholines with a high degree of unsaturation than phosphatidylcholines with low degrees of unsaturation^48^. Among our study, six of the eight identified phospholipids with polyunsaturated fatty acid chain (PC 40:5, PE 38:3, PE 38:4, PE 38:6 and PE 40:6) are replicated but also validated by a previous study^12^. This provides evidence that the associations of the specific phospholipids and blood pressure are genetically driven. *FADS1, FADS2* and *TMEM258* are all located on chromosome 11 and band q12.2 (*11q12*.*2*) in linkage disequilibrium. Our findings that they are also genome-widely significantly associated with heart and pulse rate raise the chance that phospholipids metabolism may be implicated in the relationship with blood pressure through the pleiotropic effect of genes located in *11q12*.*2*. An in-depth study in the (pleiotropic) role of gene *FADS/TMEM258* on the association of phospholipids, blood pressure and these traits is highly suggested.

The strengths of this study include the use of detailed characterized phospholipid data in a large sample size, as well as the use of replication panels. A multi-omics approach and the integration of genomic, metabolomic and epidemiologic data were performed to maximize the in-depth research of the mechanism. One of the limitations is the small number of incident hypertension cases in the current study. However, the integration of genetic data has raised an interesting hypothesis to be tested in future pathophysiological studies, in human beings and animals. To our knowledge, this is the first study performing MR on phospholipids and blood pressure. Though no significant findings were identified in the current study, the development of high-throughput technology on lipidomics will facilitate the discovery of more genetic determinants for the phospholipids and improve the strength of the instrumental variables. Previous studies reported that some anti-hypertensive drugs may have an effect on metabolism as well^49,50^. In the current study, we found a significant association of anti-hypertensive drug intake and the blood pressure related phospholipids. Following the route of the previous large GWAS study of blood pressure which adjusted for anti-hypertensive drugs intake and using MR to overcome confounders in the association of blood pressure and phospholipids, we still cannot fully exclude the effect of anti-hypertensive drugs on phospholipids. Indeed, one of the genes we identified, *ALDH1A2* has been implicated in coronary artery calcification^51,52^ and is known to interact with atenolol, a beta blocker that is prescribed to treat high blood pressure and irregular heartbeats (arrhythmia)^53^.This asks for more careful exploration of the difference between the effect of hypertension and the effect of anti-hypertensives.

In conclusion, we show eight phospholipids in the circulation that significantly associate with blood pressure and show strong clustering with components of cardiometabolic disease. These phospholipids collectively associate with incident hypertension and improve the discrimination effect of previous prediction model. Our cross-omics analyses show that phospholipid metabolites in circulation may yield insight into blood pressure regulation and raise a number of testable hypothesis for future research.

## Data Availability

The summary statistics of the meta-analysis, replication and other relevant data supporting the key findings of this study are available within the article and its supplementary information files; the cohort data sets generated and analyzed during the current study are available from the authors from each cohort upon reasonable request. No custom code or mathematical algorithm was used in the current study.

## Ethics declarations

CROATIA-VIS: All subjects were asked to provide written consent, after being informed on the study goals and main approaches, in accordance with the Declaration of Helsinki. The study was approved by the ethics committees of the University of Zagreb (No. 018057) and the University of Split School of Medicine (No. 2181-198-A3-04110-11-0008), Croatia and the Multi-Centre Research Ethics Committee for Scotland (No. 01/0/71).

ERF: The study protocol was approved by the medical ethics board of the Erasmus Medical Center Rotterdam, the Netherlands. All participants gave their informed consent in writing.

MICROS: A detailed information sheet and a form for the written informed consent were provided to each prospective participant to approve. The study was approved by the Ethics Committee of the Autonomous Province of Bolzano.

NSPHS: The NSPHS study was approved by the local ethics committee at the University of Uppsala (Regionala Etikprövningsnämnden, Uppsala, Dnr 2005:325) in compliance with the Declaration of Helsinki. All participants gave their written informed consent to the study. For participants of under legal age, a legal guardian also signed. The procedure used to obtain informed consent and the respective informed consent form has been recently discussed according to current ethical guidelines.

ORCADES: ORCADES received ethical approval from the appropriate research ethics committees in 2004. Data collection was carried out in Orkney between 2005 and 2007. Informed consent and blood samples were provided by 1019 Orcadian volunteers who had at least one grandparent from the North Isles of Orkney.

## Contributions

J.L., P.S.d.V., C.M.v.D., A. Dehghan and A. Demirkan contributed to study design. O.H.F., A.A.H., V.V., I.R., H.C. and O.P. contributed to data collection. C.H., I.R., H.C. P.P.P., J.F.W., U.G., C.M.v.D and A. Dehghan contributed to cohort design and management. J.L., P.S.d.V., F.D.G.M., A.J. and K.E.S. contributed to data analysis. J.L., P.S.d.V., K.W.v.D., C.M.v.D., A.Dihghan, A. Demirkan contributed to writing of manuscript. J.L., P.S.d.V., F.D.G.M., A.J., K.E.S., C.H., K.W.v.D., O.H.F., A.A.H., V.V., I.R., H.C., O.P., P.P.P., J.F.W., U.G., C.M.v.D., A. Dehghan and A. Demirkan contributed to critical review of manuscript.

## Acknowledgement

We would give our acknowledgement to Prof Gerd Schmitz and his team at the Institute for Clinical Chemistry and Laboratory Medicine, University Hospital Regensburg, Regensburg, Germany for generating the phospholipid data.

CROATIA-VIS: We thank a large number of individuals for their individual help in organizing, planning, and carrying out the field work and data management related to the VIS study: Professor Pavao Rudan and the staff of the Institute for Anthropological Research in Zagreb, Croatia (organization of the field work, anthropometric and physiological measurements, and DNA extraction); Professor Ariana Vorko-Jovic and the staff and medical students of the Andrija Stampar School of Public Health of the Faculty of Medicine, University of Zagreb, Croatia (questionnaires, genealogical reconstruction and data entry); Dr. Branka Salzer from the biochemistry lab “Salzer,” Croatia (measurements of biochemical traits); local general practitioners and nurses (recruitment and communication with the study population); and the employees of several other Croatian institutions who participated in the field work, including but not limited to the University of Rijeka and Split, Croatia; Croatian Institute of Public Health; Institutes of Public Health in Split and Dubrovnik, Croatia. SNP Genotyping of the Vis samples was carried out by the Genetics Core Laboratory at the Wellcome Trust Clinical Research Facility, WGH, Edinburgh.

ERF: For the ERF Study, we are grateful to all participants and their relatives, to general practitioners and neurologists for their contributions, to P. Veraart for her help in genealogy, to Jeannette Vergeer for the supervision of the laboratory work, and to P. Snijders for his help in data collection.

MICROS: For the MICROS study, we thank the primary care practitioners Raffaela Stocker, Stefan Waldner, Toni Pizzecco, Josef Plangger, Ugo Marcadent, and the personnel of the Hospital of Silandro (Department of Laboratory Medicine) for their participation and collaboration in the research project.

NSPHS: We are grateful for the contribution of district nurse Svea Hennix for data collection and Inger Jonasson for logistics and coordination of the NSPHS health survey. We would also like to thank all the participants from the community for their interest and willingness to contribute to this study.The computations were performed on resources provided by SNIC through Uppsala Multidisciplinary Center for Advanced Computational Science (UPPMAX).

ORCADES: DNA extractions for ORCADES were performed at the Wellcome Trust Clinical Research Facility in Edinburgh. We would like to acknowledge the invaluable contributions of Lorraine Anderson and the research nurses in Orkney,DNA extractions were performed at the Edinburgh Clinical Research Facility, University of Edinburgh. We would like to acknowledge the invaluable contributions of the research nurses in Orkney, the administrative team in Edinburgh and the people of Orkney.

