## Supplementary Figure for "A multi-omics study of circulating phospholipid markers of blood pressure"

**Supplementary Figures**

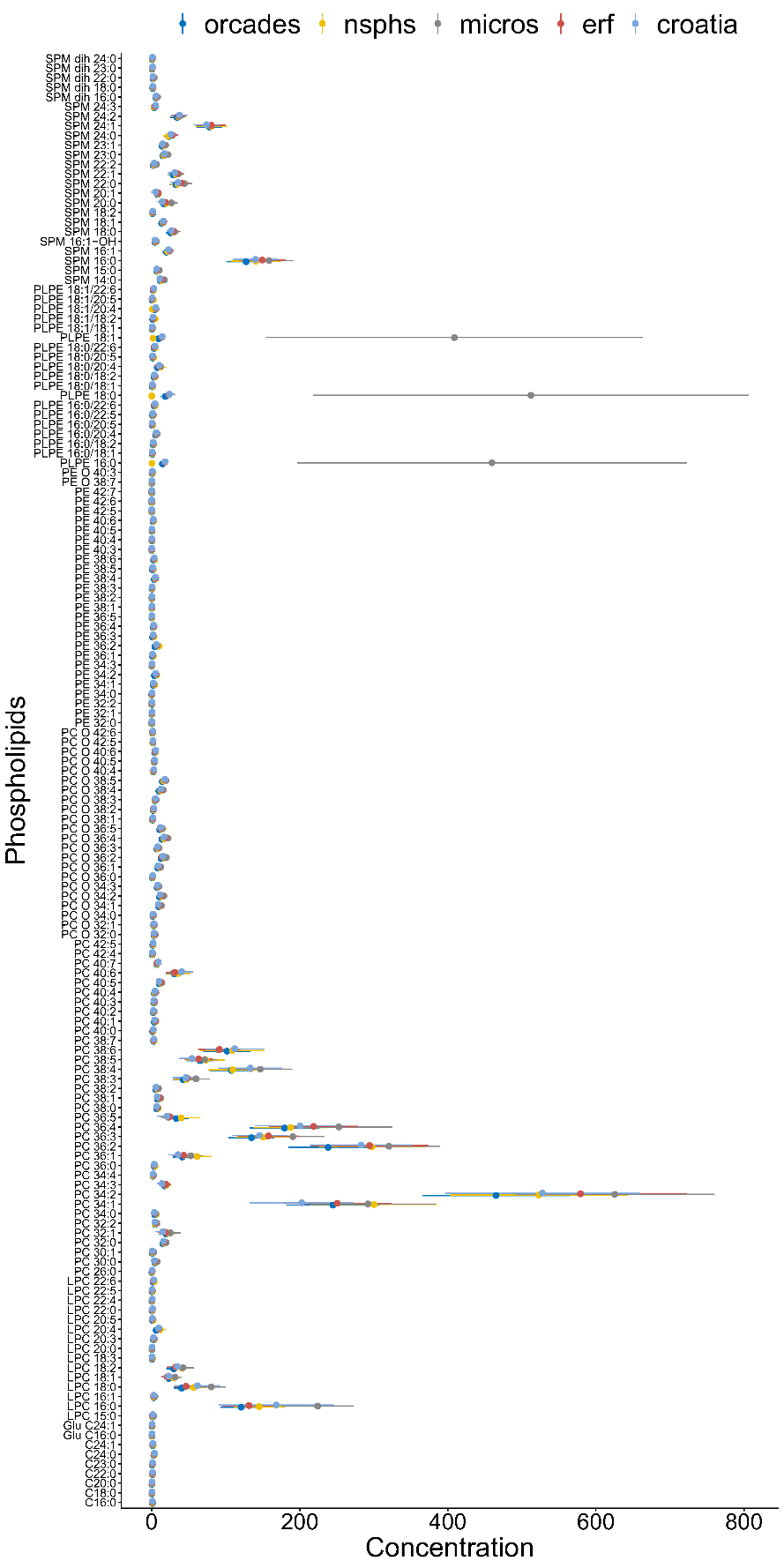

**Supplementary Figure 1** Mean and standard deviation of concentration of each phospholipid across cohorts.

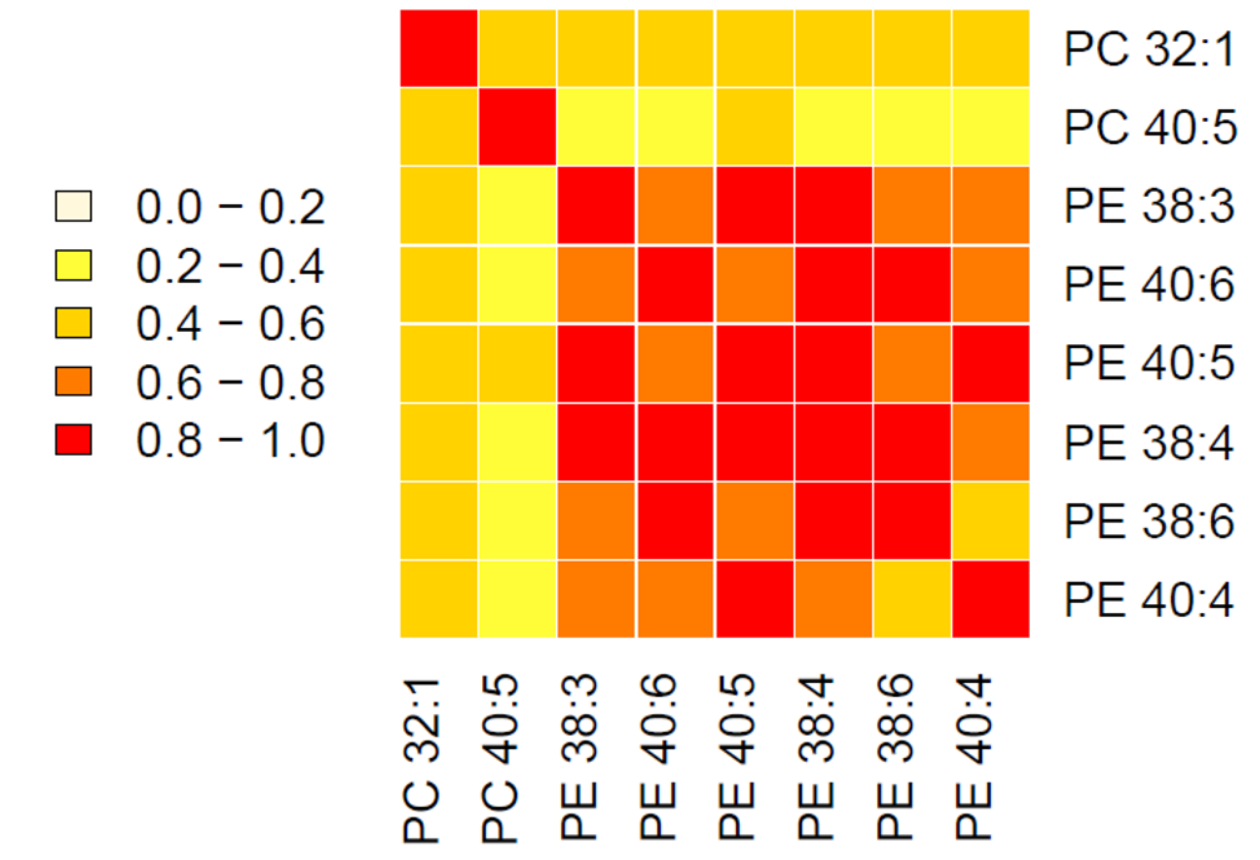

**Supplementary Figure 2 Heatmap of the correlation between the phospholipids associated with blood pressure.** The correlation was tested by Pearson's correlation in ERF (n=818).
